# Socio-economic determinants of SARS-CoV-2 infection: results from a population-based serosurvey in Geneva, Switzerland

**DOI:** 10.1101/2022.05.09.22274838

**Authors:** Hugo-Alejandro Santa-Ramírez, Ania Wisniak, Nick Pullen, María-Eugenia Zaballa, Francesco Pennacchio, Elsa Lorthe, Roxane Dumont, Hélène Baysson, Idris Guessous, Silvia Stringhini, the Specchio-COVID19 study group

## Abstract

**Background:** SARS-CoV-2 infection and its health consequences have disproportionally affected disadvantaged socio-economic groups globally. This study aimed to analyze the association between socio-economic conditions and having developed anti-SARS-CoV-2 antibodies in a population-based sample in the canton of Geneva, Switzerland.

**Methods:** Data was obtained from a population-based serosurvey of adults in Geneva and their household members, between November and December, 2020, towards the end of the second pandemic wave in the canton. Participants were tested for anti-SARS-CoV-2 antibodies. Socio-economic conditions representing different dimensions were self-reported. Mixed effects logistic regressions were conducted for each predictor to test its association with seropositive status as the main outcome.

**Results:** 2,889 adults completed the study questionnaire and were included in the final analysis. Retired participants and those living in suburban areas had lower odds of a seropositive result when compared to employed participants (OR 0.42, 95% CI - 0.20 – 0.87) and those living in urban areas (OR 0.67, 95% CI - 0.46 – 0.97), respectively. People facing financial hardship for less than a year had higher odds of a seropositive result compared to those who had never faced them (OR 2.23, 95% CI - 1.01 – 4.95). Educational level, occupational position and household income were not associated with being seropositive, nor were ethnicity or country of birth.

**Discussion:** While traditional measures of socio-economic position did not seem to be related to the risk of being infected in this sample, this study sheds lights on the importance of examining the broader social determinants of health when evaluating the differential impact of the pandemic within the population.

## 1 Introduction

Since the beginning of the COVID-19 pandemic, studies have shown that SARS-COV-2 infection and its health-related consequences have disproportionally affected disadvantaged socio-economic groups (1–3). Disadvantaged populations accumulate several vulnerabilities to infection, such as poor living conditions, higher job instability, fewer job opportunities, poorer social benefits and lower financial security (4,5), household crowding and possible impairments of their immune status due, among others, to work-related and financial stress (6). This may lead to a higher need of continued work outside the home, particularly for essential workers. Socioeconomically disadvantaged populations are also known to have a higher burden of chronic diseases and reduced access to healthcare (7), both risk factors for COVID-19 severity (8). In New York City, underprivileged neighborhoods, neighborhoods with higher household density, and those with higher proportions of black and immigrant populations were more likely to have a positive COVID-19 test result (9). An analysis of data reported to the Swiss Federal Office of Public Health (SFOPH) during the first year of the pandemic revealed that people living in neighborhoods with a low socioeconomic position index were less likely to get tested, but had a higher proportion of positive SARS-CoV-2 RT-PCR and antigen test results and were more likely to be hospitalized or die compared to people living in socioeconomically advantaged areas (10). Another study has also shown persistence of SARS-CoV-2 clusters in more disadvantaged neighborhoods, when analyzing RT-PCR positive test results (11). Several studies revealing social inequalities related to COVID-19 have been based on confirmed RT-PCR test results, therefore missing a large part of the population who did not undergo testing (12,13). Socio-economic conditions may also influence the probability of getting tested when presenting with symptoms of COVID-19. A better picture of the distribution of the infection in the population is achieved with serological surveys as they yield more accurate estimations of the real number of infections including mild and asymptomatic cases (14). Further, most studies rely on area-based indicators of socioeconomic status, thereby not allowing a more precise characterization of factors associated with SARS-CoV-2 infection.

Previous work by our research team showed associations between employment status and seropositivity during the first wave of the epidemic in the canton of Geneva, with retirees having lower odds of a seropositive result, and found no association with education, occupational status and neighborhood median income (15). A serological survey conducted among essential workers in Geneva after the first epidemic wave showed significant variation in seroprevalence across occupations (16). Nevertheless, other features that might influence serological status could not be assessed in those studies, such as ethnicity, individual income, country of birth and living and residential conditions. Although the canton of Geneva never followed a strict lockdown, there were some differences between the first and second waves, with the relaxation of certain measures such as re-opening of primary schools, as well as shops and establishments, and allowing larger social gatherings. During the second wave, a more strict use of facemasks was mandated and tests were made available free of charge to any person with symptoms.

Understanding the influence of socioeconomic conditions on the probability of being infected with SARS-CoV-2 is crucial for the implementation of equity-driven public health measures both to contain the spread of the virus during the pandemic phase and to structure the public health response in the post-pandemic phase. This study aimed to analyze the association between socioeconomic conditions and having developed anti-SARS-CoV-2 antibodies during the second COVID-19 wave (October-December 2020) in a representative sample of the population in the canton of Geneva.

## 2 Methods

The study sample included adults aged 18 years and older, who were randomly selected from a previous population-based serosurvey conducted in the canton of Geneva in spring 2020, and from population registries of the canton. Household members of recruited participants were also invited to take part in the study. Details of the selection process have been described previously (17). Recruitment occurred between November 23 and December 23, 2020. Participants were required to fill in a questionnaire (online or in paper format) and had their blood drawn to determine their SARS-CoV-2 serological status. The study was approved by the Geneva Cantonal Commission for Research Ethics (Project N° 2020-00881). All participants provided informed written consent.

Socio-economic conditions were assessed through conventional indicators of socio-economic status, such as self-reported occupational position, education, family income, and other social determinants, including ethnicity, country of birth, household residential area, household density and the experience of financial hardship. Detailed information on the variables used is available in the supplementary material (Annex I). Serological status was determined using the Elecsys anti-SARS-CoV-2 S assay (Roche Diagnostics, Rotkreuz, Switzerland) detecting total immunoglobulins (IgM/A/G) targeting the spike protein, following manufacturer’s recommendations (≥0·8 U/mL considered seropositive). Of note, the vaccination campaign in Switzerland started on December 23th, 2020. Thus, antibodies detected during this study could only have been produced in response to a SARS-CoV-2 infection.

Mixed effects logistic regressions were conducted for each individual predictor with seropositive status as the main outcome and the household as the second level random effect variable. Four types of models were developed: one model adjusted for age and sex only, another model additionally adjusted for education, occupational position and family income; a third model adjusted for health-related variables (weight status classified through categories of BMI, having a chronic disease, smoking status and blood group); and a final model adjusted for all of the variables used in the previous models (Annex II. Supplementary material). Reference categories were set to the most socially advantaged groups. Multicollinearity was assessed for each of the adjusted models with no variables showing noticeable collinearity. Analyses were conducted in the overall population and stratified by sex, as a differential risk for COVID-19 outcomes and SARS-CoV-2 infection have been documented between men and women (19) (Annex III and IV. Supplementary material). Results have not been adjusted for multiple comparisons. Statistical analyses were conducted using STATA version 14.0 (StataCorp, College Station, TX, USA).

## 3 Results

A total of 2,986 adults participated in the study and had a blood sample taken, of which 2,889 completed the study questionnaire and were included in the final analysis. The mean (SD) age of participants was 47.8 (15.4) years, and 55% were women. Education, occupation and income were not associated with being seropositive in the overall sample (Table 1 and Annex II. Supplementary material). Looking at other socioeconomic indicators, associations were found with employment status, financial hardship and the residential area in the overall sample, with retired people and those living in a suburban area exhibiting lower odds of a seropositive result when compared with those employed and those living in an urban area, respectively. People facing financial hardship for less than a year had twice the odds of a seropositive result when compared to those that had never faced financial difficulties, all other variables remaining constant. This association did not hold for participants having faced financial difficulties for several years. People living in households with higher density also tended to have higher odds of a seropositive result. Ethnicity and country of birth were not associated with seropositivity in our sample. When stratifying by sex, men in the lower occupational position tended to have higher odds of a seropositive result when compared to those with a higher occupational position (OR 1.79, 95% CI - 0.97, 3.32) (Annex IV. Supplementary material). Higher odds of a seropositive result were found for unemployed women compared to employed women (OR 2.01, 95% CI - 1.01, 4.03) (Annex III. Supplementary material).

**Table 1.**
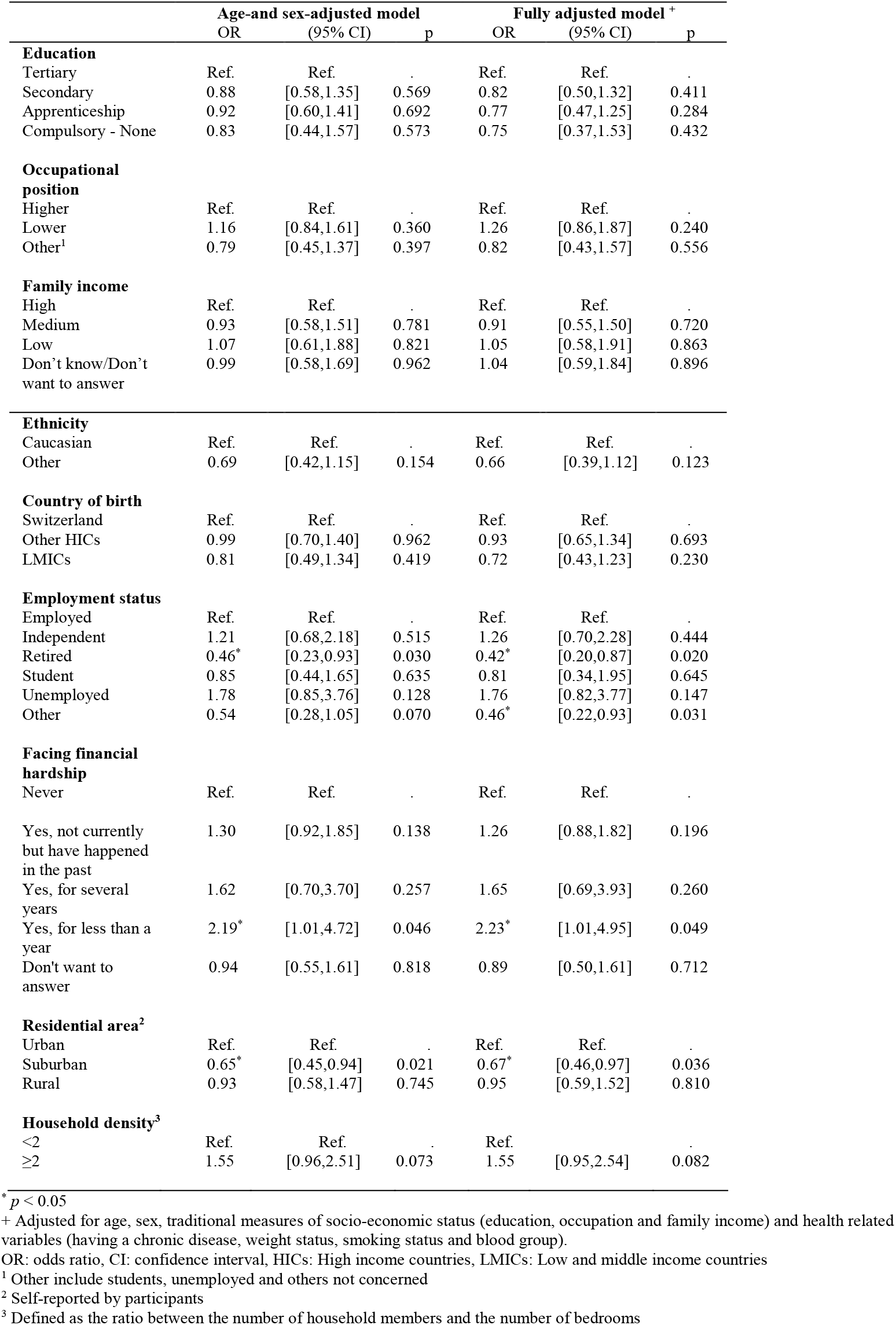
Association between socio-economic predictors and seropositive status to SARS-CoV-2 in the overall population.

## 4 Discussion

In this population-based serological study, we found associations between financial hardship, employment status, residential area and the odds of having developed anti-SARS-CoV-2 antibodies. A higher household density tended to be associated with increased odds of a seropositive result. However, other socioeconomic conditions such as educational level, occupational position and income were not associated with serological status, nor were ethnicity and country of birth.

Our study shows lower odds of a SARS-CoV-2 seropositive result for the retired population when compared with the employed one, possibly due to the fact of being considered at higher risk of severe forms of COVID-19, potentially leading them to reduce social contact and increase the use of preventive measures. This result is consistent with previous findings from the first seroprevalence study in Geneva (15) and findings from a report in the UK for the age group comprising the retired population (20).

We also found a protective effect of the residential area for people living in suburban areas compared to urban areas, which could be explained by increased use of private transportation and lower population density. While this may also be the case in rural areas, higher commuting times and a potentially lower sense of danger posed by the infection in these areas may explain the lack of significant difference in seropositivity between rural and urban areas. It has been suggested that a lower population density outside the urban areas might have contributed to lower incidence at the beginning of the pandemic in some regions in Europe (21) and some studies have shown lower seroprevalence in municipalities of less than 100.000 inhabitants (22). Further work is needed to uncover the potential mechanisms explaining the association of the residential area with a seropositive result in the population of Geneva, as considering the small size of the canton, the difference between urban and suburban areas is not clearly established and the distribution of SARS-CoV-2 infections might not follow a similar pattern as the one found in other places.

There seemed to be a trend in the association between duration of financial difficulties and the odds of seropositivity, with people facing financial hardship for less than one year having the highest odds of a seropositive result compared to those who reported never facing financial hardship. This could potentially be explained by the development of coping mechanisms in individuals being used to financial difficulties, while those with unexpected economic hardship may need more time to adapt to their new circumstances, putting them at higher risk of SARS-CoV-2 exposure as they cannot afford to miss work or need to look for economic alternatives. A consistent association of financial hardship due to COVID-19 with health behavior risk changes has been shown in a sample of women in the U.S., although the health behaviors assessed were based on lifestyle factors rather than on the risk of getting infected with SARS-CoV-2 (23); this may support a hypothesis of higher risky behaviors when facing economic stress. On the other hand, reverse causation cannot be excluded, with people affected by COVID-19 being more likely to reduce their work time due to symptomatic disease leading to financial instability.

Consistent with our previous findings (15), we did not find associations between educational level, occupational position, income, ethnicity or country of birth and the seropositive status. Other studies conducted in European countries have found similar results for education (20,24). However, there is divergence when looking at the role of occupation, income, nationality and ethnicity, with studies showing conflicting results (20,25–27). This may be due to differences in survey design and measurement across studies. The association between education and seropositivity may be confounded by increased SARS-CoV-2 exposure in certain professions requiring tertiary education, such as in the health-related field. Future analyses should take into account professional exposure to SARS-CoV-2. Heterogeneity in the socioeconomic circumstances in different countries, as well as diverging policies for pandemic management, may also explain some of the conflicting results.

Strengths of this study include the relatively large sample size and comprehensive information related to different social and economic circumstances at the individual level as well as objective information about individual health such as the serological status. Our study also has some limitations. A selection bias should not be disregarded, with people with higher health concerns being more prone to participate, and those most socioeconomically disadvantaged less likely to be included, limiting the generalizability of our results. In addition, the population that was hospitalized at the time of the study or that died because of COVID-19 could not be included in the study, therefore potentially masking the association between socio-economic conditions and SARS-CoV-2 seropositivity for severe cases. As other studies have documented, the severity of the disease might be higher in socioeconomically disadvantaged groups (4,28).

The COVID-19 pandemic has disproportionately affected socially vulnerable populations globally. However the impact of socio-economic determinants can vary widely depending on geographical, political and cultural contexts (29–31). In our study we have found associations of employment status, financial hardship and residential area with the natural development of anti SARS-CoV-2 antibodies during the second wave of the pandemic (before the roll-out of the vaccination campaign in Switzerland); but not with other socioeconomic conditions. Our results highlight the importance of examining the broader social determinants of health when evaluating the differential impact of the pandemic within the population. A better understanding of the structural determinants shaping the inequitable distribution of COVID-19 among the population is imperative for tailoring public health interventions, such as vaccine prioritization and public health campaigns, and for setting up supportive mechanisms for vulnerable population groups.

## Data Availability

Data produced in the present study are available upon reasonable request to the corresponding author

## 6 Author Contributions

SS and IG conceived the study. AW, NP, HB, MZ, FP, RD and SS contributed to the scientific coordination and data management during the study. HS and AW drafted the first version of the manuscript. HS and NP did data analyses. NP, EL, HB, MZ, FP, IG and SS contributed to draft the manuscript. All authors reviewed and approved the final manuscript.

### For the Specchio-COVID19 study group

Deborah Amrein, Isabelle Arm-Vernez, Andrew S Azman, Fatim Ba,, Delphine Bachmann, Antoine Bal, Jean-François Balavoine, Michael Balavoine, Rémy P Barbe, Hélène Baysson, Lison Beigbeder, Julie Berthelot, Patrick Bleich, Livia Boehm, Gaëlle Bryand, Viola Bucolli, François Chappuis, Prune Collombet, Delphine Courvoisier, Alain Cudet, Vladimir Davidovic, Carlos de Mestral Vargas, Paola D’ippolito, Richard Dubos, Roxane Dumont, Isabella Eckerle, Nacira El Merjani, Antoine Flahault, Natalie Francioli, Marion Frangville, Clément Graindorge, Idris Guessous, Séverine Harnal, Samia Hurst, Laurent Kaiser, Omar Kherad, Julien Lamour, Pierre Lescuyer, François L’Huissier, Fanny-Blanche Lombard, Andrea Jutta Loizeau, Elsa Lorthe, Chantal Martinez, Lucie Ménard, Lakshmi Menon, Ludovic Metral-Boffod, Benjamin Meyer, Alexandre Moulin, Mayssam Nehme, Natacha Noël, Francesco Pennacchio, Javier Perez-Saez, Didier Pittet, Jane Portier, Klara M Posfay-Barbe, Géraldine Poulain, Caroline Pugin, Nick Pullen, Zo Francia Randrianandrasana,, Viviane Richard, Frederic Rinaldi, Jessica Rizzo, Deborah Rochat, Irine Sakvarelidze, Khadija Samir, Santa Ramirez Hugo Alejandro, Stephanie Schrempft, Claire Semaani, Silvia Stringhini, Stéphanie Testini, Deborah Urrutia Rivas, Charlotte Verolet, Jennifer Villers, Guillemette Violot, Nicolas Vuilleumier, Ania Wisniak, Sabine Yerly, María-Eugenia Zaballa

## 7 Data Availability Statement

## 9 Acknowledgments

We thank all the participants, without whom this study would not have been possible

## 11 Supplementary Material

The supplementary material for this article can be found in the additional document

