## Supplementary material for "Socio-economic determinants of SARS-CoV-2 infection: results from a population-based serosurvey in Geneva, Switzerland"

### 1 Supplementary Data

#### Annex I. Operationalization of variables and demographic characteristics of the study population

#### Operational definitions

Occupational position was categorized based on the European Socioeconomic Classification (ESEC)[i] in Professional/Managers (I) (ESEC 1-2), Higher grade White Collar workers (II) (ESEC 3), independent (III) (ESEC 4-5), Lower grade white collar workers (IV) (ESEC 7), Blue collar workers (V) (ESEC 6, 8, 9) and other (VI) (including students, unemployed and others non concerned). For conducting the final models, occupational position was further categorized into higher position (I – III), lower position (IV and V) and other (VI).

Education was categorized as tertiary (doctoral, university, superior professional training and other training accounting for tertiary degree diplomas), secondary (secondary school), apprenticeships (certified and non-certified training), and compulsory/none.

Family income was categorized using the tables for household income according to the type of household of the Cantonal Office of Statistics of Geneva for 2015-2017[ii]. The type of households taken into consideration included couples with children, couples without children, single-parent households, one-person households and households of several people. A threshold was set for each type of household as low when below the quartile 1 of the income distribution, medium in between quartile 1 and 3, and high above quartile 3. The participants' family income was classified into each of the categories according to the type of household they belonged to. A fourth category accounting for don't know/no answer was included due to a high proportion of responses.

Ethnicity was categorized as Caucasian or other, given the low number of participants reporting each of the options other than Caucasian in the questionnaire. Other included people with Asian, African, Arabic, Latin American and the Indian Subcontinent origin, as well as those who did not want to answer. Country of birth was classified as Switzerland, other High income countries (HICs), or Low and Middle income countries (LMICs), according to the World Bank income economies classification for the fiscal year 2021[iii].

Employment status was classified as employed, independent, retired, student, unemployed, and other (including household activities and people with disabilities).

Financial hardship was categorized as those who have never faced it and those facing it currently or in the past. Those who have faced financial hardship were further categorized according to the duration of financial hardship in: not currently but has happened in the past, facing it for less than one year, and facing it for several years.

Variables related to the living environment included the residential area and household density. The former was classified as urban, suburban (outskirts of a city) or rural area based on the self-reported answer of participants. The latter was created as the ratio of persons in the household divided by the number of bedrooms of their house, setting a threshold of 2.

Health-related variables in this analysis included having a chronic disease, weight status, smoking status and blood group. Weight status was categorized as normal weight, underweight and overweight/obese according to WHO categories of BMI. Smoking status was categorized as never or former smokers and current smokers. Blood group was classified as O, other group and don't know, as some studies showed a protective effect of blood group O to SARS-CoV-2 infection compared to others. Having a history of close contact with a SARS-CoV-2 infected person was independently assessed to validate the results as it has been previously shown to have a strong association with seropositive status[iv].

### Description of the study sample

From a total of 2,986 adults participating in the study, 2,889 participants (55% women) had complete information on the variables of interest and were considered for final analyses. The mean age of participants was 47.8 ± 15.4 years. Most of the study population were of European-Caucasian origin (88.75%), with a higher proportion than the one expected for the general population, nonetheless the majority of the resident population in the Canton is also from European origin (73%)[v]. Our study population had a higher proportion of people with secondary or tertiary education (76.1%). The distribution of education in the population of Geneva has the highest proportion of people with tertiary degrees (41%), followed by those with secondary degrees (39%)[vi]. Similar results are observed for the distribution of occupation categories in our study, with a higher proportion of professional or managers, followed by high grade white collar workers, which may be in line with the majority of the population in Geneva working in the tertiary sector (87.5%)[vii]. As for the employment status, employed people accounted for the highest proportion (58%) followed by retirees (16.6%). Half of the participants lived within a city and a low proportion of the population reported currently experiencing financial hardship (7.4%) (Supplementary table 1).

Regarding the health status of participants, 35.1% had excess weight and 23.8% reported having a chronic disease. 17.8% of participants reported being current smokers, a lower proportion than the one found for the general population of the canton (27.1%)[viii]. In our study, 32% of participants reported having O blood group, showing a similar pattern for the distribution of the blood groups to the one found in the general population, with the highest proportion of people belonging to the group O, followed by group A[ix] (Supplementary table 2).

When accounting for the effects of health-related variables, higher odds of a seropositive result were found for people being overweight or obese and for those with other blood groups compared to the group O (Annex II, II and IV). As shown in previous work[iv], there was a strong association of having a confirmed COVID-19 positive contact with seropositive status (Annex II, III and IV).

### 2 Supplementary Figures and Tables

**Supplementary table 1.** Demographic and socio-economic characteristics of study participants

| | n | % |
|---|---|---|
| **Seropositive status** | | |
| Negative | 2306 | 79.5 |
| Positive | 593 | 20.5 |
| **Sex** | | |
| Women | 1594 | 55 |
| Men | 1305 | 45 |
| **Age** | | |
| 18 – 34 | 523 | 18 |
| 35 – 49 | 1201 | 41.4 |
| 50 – 64 | 708 | 24.4 |
| ≥65 | 467 | 16.1 |
| **Education** | | |
| Tertiary | 1783 | 60.2 |
| Secondary | 460 | 15.9 |
| Apprenticeship | 498 | 17.2 |
| Compulsory/None | 193 | 6.7 |
| **Occupational category** | | |
| Professional/Manager | 891 | 30.8 |
| High grade white collar | 646 | 22.4 |
| Independent | 32 | 1.1 |
| Low grade white collar | 693 | 24 |
| Blue collar | 372 | 12.9 |
| Other | 256 | 8.9 |
| **Family income** | | |
| High | 391 | 13.5 |
| Medium | 1383 | 47.8 |
| Low | 502 | 17.3 |
| Don’t know/ Don't want to answer | 619 | 21.4 |
| **Ethnicity** | | |
| Caucasian | 2562 | 88.7 |
| Other | 325 | 11.3 |
| **Country of birth** | | |
| Switzerland | 1655 | 57.2 |
| Other HICs | 905 | 31.3 |
| LMICs | 333 | 11.5 |
| **Employment status** | | |
| Employed | 1683 | 58.1 |
| Independent | 212 | 7.3 |
| Retired | 480 | 16.6 |
| Student | 224 | 7.7 |
| Unemployed | 115 | 4 |
| Other | 184 | 6.3 |

| | | |
|---|---|---|
| **Financial difficulties** | | |
| Never | 1573 | 54.3 |
| Yes, not currently but have happened in the past | 837 | 28.9 |
| Yes, for several years | 103 | 3.6 |
| Yes, for less than a year | 111 | 3.8 |
| Don't want to answer | 273 | 9.4 |
| **Residential area** | | |
| Urban | 1451 | 50.1 |
| Suburban | 978 | 33.8 |
| Rural | 467 | 16.1 |
| **Household density** | | |
| <2 | 2,147 | 84.4 |
| ≥2 | 397 | 15.6 |

**Supplementary table 2.** Health related characteristics of study participants

| | **n** | **%** |
|---|---|---|
| **SARS-CoV-2 close contact** | | |
| No | 2129 | 73.5 |
| Yes | 767 | 26.5 |
| **Weight status** | | |
| Normal weight | 1757 | 60.6 |
| Underweight | 95 | 3.3 |
| Overweight | 798 | 27.5 |
| Obese | 249 | 8.6 |
| **Chronic disease** | | |
| No | 2208 | 76.2 |
| Yes | 689 | 23.8 |
| **Smoking status** | | |
| Never smokers | 1561 | 53.9 |
| Former smokers | 820 | 28.3 |
| Current smokers | 517 | 17.8 |
| **Blood group** | | |
| A | 880 | 30.4 |
| B | 200 | 6.93 |
| AB | 104 | 3.6 |
| O | 939 | 32.5 |
| Don’t know | 768 | 26.6 |

**Annex II.** Association between socio-economic and health predictors with SARS-CoV-2 seropositive status, complete models in the overall population

| | (Model 1)<br>Demographic covariates | | | (Model 2)<br>Health covariates | | | (Model 3)<br>SES covariates | | | (Model 4)<br>Fully adjusted model | | |
|---|---|---|---|---|---|---|---|---|---|---|---|---|
| | OR | (95% CI) | p | OR | (95% CI) | p | OR | (95% CI) | p | OR | (95% CI) | p |
| **Education[±]** | | | | | | | | | | | | |
| Tertiary | Ref. | Ref. | . | Ref. | Ref. | . | Ref. | Ref. | . | Ref. | Ref. | . |
| Secondary | 0.88 | [0.58,1.35] | 0.569 | 0.86 | [0.56,1.33] | 0.496 | 0.83 | [0.52,1.34] | 0.444 | 0.82 | [0.50,1.32] | 0.411 |
| Apprenticeship | 0.92 | [0.60,1.41] | 0.692 | 0.86 | [0.56,1.34] | 0.514 | 0.79 | [0.49,1.28] | 0.335 | 0.77 | [0.47,1.25] | 0.284 |
| Compulsory - None | 0.83 | [0.44,1.57] | 0.573 | 0.83 | [0.44,1.57] | 0.562 | 0.75 | [0.38,1.51] | 0.424 | 0.75 | [0.37,1.53] | 0.432 |
| **Occupational position[±]** | | | | | | | | | | | | |
| Higher | Ref. | Ref. | . | Ref. | Ref. | . | Ref. | Ref. | . | Ref. | Ref. | . |
| Lower | 1.16 | [0.84,1.61] | 0.360 | 1.13 | [0.81,1.57] | 0.472 | 1.28 | [0.87,1.89] | 0.206 | 1.26 | [0.86,1.87] | 0.240 |
| Other | 0.79 | [0.45,1.37] | 0.397 | 0.78 | [0.44,1.37] | 0.384 | 0.82 | [0.44,1.55] | 0.544 | 0.82 | [0.43,1.57] | 0.556 |
| **Family income[±]** | | | | | | | | | | | | |
| High | Ref. | Ref. | . | Ref. | Ref. | . | Ref. | Ref. | . | Ref. | Ref. | . |
| Medium | 0.93 | [0.58,1.51] | 0.781 | 0.93 | [0.57,1.51] | 0.758 | 0.92 | [0.56,1.50] | 0.728 | 0.91 | [0.55,1.50] | 0.720 |
| Low | 1.07 | [0.61,1.88] | 0.821 | 1.07 | [0.60,1.90] | 0.813 | 1.04 | [0.58,1.88] | 0.894 | 1.05 | [0.58,1.91] | 0.863 |
| Don't know/No answer/ | 0.99 | [0.58,1.69] | 0.962 | 0.97 | [0.56,1.68] | 0.921 | 1.05 | [0.60,1.84] | 0.868 | 1.04 | [0.59,1.84] | 0.896 |
| **Ethnicity** | | | | | | | | | | | | |
| Caucasian | Ref. | Ref. | . | Ref. | Ref. | . | Ref. | Ref. | . | Ref. | Ref. | . |
| Other | 0.69 | [0.42,1.15] | 0.154 | 0.69 | [0.42,1.15] | 0.212 | 0.67 | [0.40,1.12] | 0.127 | 0.69 | [0.41,1.17] | 0.170 |
| **Country of birth** | | | | | | | | | | | | |
| Switzerland | Ref. | Ref. | . | Ref. | Ref. | . | Ref. | Ref. | . | Ref. | Ref. | . |
| Other HICs | 0.99 | [0.70,1.40] | 0.962 | 0.99 | [0.70,1.41] | 0.968 | 0.93 | [0.65,1.34] | 0.713 | 0.94 | [0.65,1.36] | 0.746 |
| LMICs | 0.81 | [0.49,1.34] | 0.419 | 0.79 | [0.48,1.33] | 0.448 | 0.75 | [0.45,1.27] | 0.282 | 0.75 | [0.44,1.27] | 0.279 |
| **Employment status** | | | | | | | | | | | | |
| Employed | Ref. | Ref. | . | Ref. | Ref. | . | Ref. | Ref. | . | Ref. | Ref. | . |
| Independent | 1.21 | [0.68,2.18] | 0.515 | 1.28 | [0.71,2.31] | 0.401 | 1.20 | [0.67,2.17] | 0.534 | 1.27 | [0.70,2.29] | 0.438 |
| Retired | 0.46[*] | [0.23,0.93] | 0.030 | 0.46[*] | [0.23,0.92] | 0.028 | 0.44[*] | [0.21,0.90] | 0.024 | 0.42[*] | [0.20,0.87] | 0.020 |
| Student | 0.85 | [0.44,1.65] | 0.635 | 0.86 | [0.44,1.69] | 0.674 | 0.80 | [0.34,1.89] | 0.613 | 0.82 | [0.34,1.98] | 0.663 |
| Unemployed | 1.78 | [0.85,3.76] | 0.128 | 1.82 | [0.86,3.87] | 0.129 | 1.72 | [0.81,3.66] | 0.156 | 1.74 | [0.81,3.73] | 0.156 |
| Other | 0.54 | [0.28,1.05] | 0.070 | 0.51 | [0.26,1.01] | 0.052 | 0.48[*] | [0.24,0.97] | 0.041 | 0.46[*] | [0.22,0.93] | 0.031 |
| **Facing financial hardship** | | | | | | | | | | | | |
| Never | Ref. | Ref. | . | Ref. | Ref. | . | Ref. | Ref. | . | Ref. | Ref. | . |
| Currently or in the past[Ƭ] | 1.40[*] | [1.01,1.95] | 0.042 | 1.38 | [0.99,1.93] | 0.056 | 1.42[*] | [1.01,1.99] | 0.041 | 1.37 | [0.97,1.93] | 0.071 |
| Yes, not currently but have happened in the past | 1.30 | [0.92,1.85] | 0.138 | 1.27 | [0.89,1.81] | 0.188 | 1.30 | [0.91,1.86] | 0.148 | 1.26 | [0.88,1.81] | 0.206 |

| | | | | | | | | | | | | |
|---|---|---|---|---|---|---|---|---|---|---|---|---|
| Yes, for several years | 1.62 | [0.70,3.70] | 0.257 | 1.69 | [0.72,3.92] | 0.226 | 1.59 | [0.68,3.75] | 0.288 | 1.69 | [0.71,4.03] | 0.239 |
| Yes, for less than a year | 2.19* | [1.01,4.72] | 0.046 | 2.22* | [1.01,4.87] | 0.046 | 2.24* | [1.02,4.91] | 0.045 | 2.25* | [1.01,5.00] | 0.047 |
| Don't want to answer | 0.94 | [0.55,1.62] | 0.831 | 0.91 | [0.53,1.58] | 0.745 | 0.93 | [0.52,1.67] | 0.802 | 0.92 | [0.51,1.67] | 0.784 |
| **Residential area** | | | | | | | | | | | | |
| Urban | Ref. | Ref. | . | Ref. | Ref. | . | Ref. | Ref. | . | Ref. | Ref. | . |
| Suburban | 0.65* | [0.45,0.94] | 0.021 | 0.65* | [0.45,0.94] | 0.021 | 0.68* | [0.47,0.98] | 0.040 | 0.67* | [0.46,0.97] | 0.036 |
| Rural | 0.93 | [0.58,1.47] | 0.745 | 0.91 | [0.57,1.46] | 0.706 | 0.97 | [0.61,1.55] | 0.894 | 0.94 | [0.59,1.52] | 0.810 |
| **Household density** | | | | | | | | | | | | |
| <2 | Ref. | Ref. | . | Ref. | Ref. | . | Ref. | Ref. | . | Ref. | Ref. | . |
| ≥2 | 1.55 | [0.96,2.51] | 0.07 | 1.56 | [0.96,2.53] | 0.072 | 1.53 | [0.94,2.50] | 0.085 | 1.55 | [0.95,2.54] | 0.082 |

| | **(Model 1) Demographic covariates** | | | **(Model 2) Health covariates** | | | **(Model 3) SES covariates** | | | **(Model 4) Fully adjusted model** | | |
|---|---|---|---|---|---|---|---|---|---|---|---|---|
| | OR | (95% CI) | p | OR | (95% CI) | p | OR | (95% CI) | p | OR | (95% CI) | p |
| **SARS-CoV-2 close contact** | | | | | | | | | | | | |
| No | Ref. | Ref. | . | Ref. | Ref. | . | Ref. | Ref. | . | Ref. | Ref. | . |
| Yes | 3.79*** | [2.90,4.96] | <0.001 | 3.80*** | [2.90,4.99] | <0.001 | 3.84*** | [2.93,5.03] | <0.001 | 3.83*** | [2.92,5.03] | <0.001 |
| **Weight status±±** | | | | | | | | | | | | |
| Normal weight | Ref. | Ref. | . | Ref. | Ref. | . | Ref. | Ref. | . | Ref. | Ref. | . |
| Underweight | 1.17 | [0.51,2.68] | 0.72 | 1.19 | [0.51,2.74] | 0.687 | 1.22 | [0.53,2.85] | 0.640 | 1.25 | [0.53,2.91] | 0.613 |
| Overweight-Obese | 1.60** | [1.15,2.24] | 0.006 | 1.57** | [1.12,2.19] | 0.009 | 1.57** | [1.12,2.20] | 0.009 | 1.54* | [1.10,2.16] | 0.013 |
| **Chronic disease±±** | | | | | | | | | | | | |
| No | Ref. | Ref. | . | Ref. | Ref. | . | Ref. | Ref. | . | Ref. | Ref. | . |
| Yes | 1.13 | [0.78,1.64] | 0.517 | 1.08 | [0.74,1.58] | 0.673 | 1.16 | [0.79,1.69] | 0.448 | 1.11 | [0.76,1.62] | 0.597 |
| **Smoking status±±** | | | | | | | | | | | | |
| Never & Former smokers | Ref. | Ref. | . | Ref. | Ref. | . | Ref. | Ref. | . | Ref. | Ref. | . |
| Current smokers | 0.80 | [0.53,1.19] | 0.268 | 0.80 | [0.53,1.20] | 0.289 | 0.80 | [0.53,1.20] | 0.281 | 0.81 | [0.53,1.22] | 0.304 |
| **Blood group±±** | | | | | | | | | | | | |
| O | Ref. | Ref. | . | Ref. | Ref. | . | Ref. | Ref. | . | Ref. | Ref. | . |
| Other | 1.47* | [1.03,2.11] | 0.035 | 1.46* | [1.02,2.09] | 0.040 | 1.44 | [1.00,2.07] | 0.050 | 1.43 | [0.99,2.05] | 0.056 |
| Don't know | 1.33 | [0.89,1.98] | 0.169 | 1.32 | [0.88,1.98] | 0.174 | 1.32 | [0.88,1.97] | 0.186 | 1.30 | [0.86,1.95] | 0.211 |

OR: odds ratio, CI: confidence interval
* $p < 0.05$, ** $p < 0.01$, *** $p < 0.001$
Model 1: adjusted for age and sex only
Model 2: adjusted for age, sex, weight status, chronic disease, smoking status and blood group
Model 3: adjusted for age, sex, education, occupational position and family income
Model 4: adjusted for all the above
± Models 3 and 4 adjusted for the other SES covariates
±± Models 2 and 4 adjusted for the other health covariates
Ʈ An aggregated analysis with the collapsed categories was conducted first and further disaggregated in the categories below

**Annex III.** Association between socio-economic and health predictors with SARS-CoV-2 seropositive status, complete models in women

| | (Model 1) | | | (Model 2) | | | (Model 3) | | | (Model 4) | | |
|---|---|---|---|---|---|---|---|---|---|---|---|---|
| | Demographic covariates | | | Health covariates | | | SES covariates | | | Fully adjusted model | | |
| | OR | (95% CI) | p | OR | (95% CI) | p | OR | (95% CI) | p | OR | (95% CI) | p |
| **Education[±]** | | | | | | | | | | | | |
| Tertiary | Ref. | Ref. | . | Ref. | Ref. | . | Ref. | Ref. | . | Ref. | Ref. | . |
| Secondary | 0.91 | [0.63,1.32] | 0.628 | 0.91 | [0.63,1.32] | 0.638 | 0.96 | [0.63,1.46] | 0.831 | 0.96 | [0.63,1.46] | 0.850 |
| Apprenticeship | 1.03 | [0.69,1.53] | 0.900 | 1.04 | [0.69,1.54] | 0.864 | 1.09 | [0.69,1.70] | 0.723 | 1.09 | [0.70,1.71] | 0.700 |
| Compulsory - None | 0.77 | [0.43,1.36] | 0.367 | 0.79 | [0.45,1.41] | 0.432 | 0.80 | [0.43,1.50] | 0.487 | 0.83 | [0.44,1.55] | 0.554 |
| **Occupational position[±]** | | | | | | | | | | | | |
| Higher | Ref. | Ref. | . | Ref. | Ref. | . | Ref. | Ref. | . | Ref. | Ref. | . |
| Lower | 0.91 | [0.67,1.22] | 0.516 | 0.92 | [0.68,1.24] | 0.569 | 0.93 | [0.65,1.32] | 0.671 | 0.93 | [0.65,1.33] | 0.690 |
| Other | 0.96 | [0.59,1.57] | 0.882 | 0.95 | [0.59,1.54] | 0.839 | 0.97 | [0.56,1.67] | 0.902 | 0.94 | [0.55,1.63] | 0.838 |
| **Family income[±]** | | | | | | | | | | | | |
| High | Ref. | Ref. | . | Ref. | Ref. | . | Ref. | Ref. | . | Ref. | Ref. | . |
| Medium | 0.81 | [0.52,1.27] | 0.368 | 0.81 | [0.52,1.26] | 0.351 | 0.83 | [0.53,1.30] | 0.410 | 0.82 | [0.52,1.28] | 0.380 |
| Low | 0.94 | [0.56,1.58] | 0.824 | 0.95 | [0.57,1.59] | 0.844 | 0.96 | [0.57,1.63] | 0.883 | 0.96 | [0.57,1.63] | 0.876 |
| Don't know/No answer/ | 0.84 | [0.52,1.36] | 0.479 | 0.84 | [0.51,1.36] | 0.474 | 0.88 | [0.53,1.45] | 0.609 | 0.87 | [0.53,1.44] | 0.588 |
| **Ethnicity** | | | | | | | | | | | | |
| Caucasian | Ref. | Ref. | . | Ref. | Ref. | . | Ref. | Ref. | . | Ref. | Ref. | . |
| Other | 0.63 | [0.39,1.01] | 0.056 | 0.64 | [0.40,1.02] | 0.061 | 0.64 | [0.39,1.03] | 0.068 | 0.64 | [0.40,1.04] | 0.071 |
| **Country of birth** | | | | | | | | | | | | |
| Switzerland | Ref. | Ref. | . | Ref. | Ref. | . | Ref. | Ref. | . | Ref. | Ref. | . |
| Other HICs | 0.97 | [0.71,1.32] | 0.834 | 0.96 | [0.70,1.31] | 0.807 | 0.94 | [0.68,1.31] | 0.719 | 0.93 | [0.68,1.29] | 0.680 |
| LMICs | 0.80 | [0.51,1.25] | 0.322 | 0.79 | [0.51,1.23] | 0.301 | 0.80 | [0.51,1.27] | 0.342 | 0.79 | [0.50,1.24] | 0.303 |
| **Employment status** | | | | | | | | | | | | |
| Employed | Ref. | Ref. | . | Ref. | Ref. | . | Ref. | Ref. | . | Ref. | Ref. | . |
| Independent | 1.26 | [0.73,2.18] | 0.408 | 1.26 | [0.73,2.19] | 0.392 | 1.23 | [0.71,2.12] | 0.463 | 1.23 | [0.72,2.13] | 0.449 |
| Retired | 0.70 | [0.36,1.34] | 0.281 | 0.66 | [0.34,1.27] | 0.216 | 0.69 | [0.35,1.34] | 0.273 | 0.65 | [0.33,1.28] | 0.212 |
| Student | 1.23 | [0.68,2.23] | 0.492 | 1.23 | [0.68,2.22] | 0.488 | 1.22 | [0.57,2.59] | 0.611 | 1.21 | [0.57,2.57] | 0.613 |
| Unemployed | 2.06* | [1.02,4.13] | 0.043 | 2.00* | [1.00,4.01] | 0.049 | 2.07* | [1.03,4.15] | 0.041 | 2.02* | [1.01,4.03] | 0.047 |
| Other | 0.85 | [0.51,1.39] | 0.513 | 0.81 | [0.49,1.32] | 0.397 | 0.81 | [0.48,1.37] | 0.425 | 0.77 | [0.46,1.30] | 0.331 |
| **Facing financial hardship** | | | | | | | | | | | | |
| Never | Ref. | Ref. | . | Ref. | Ref. | . | Ref. | Ref. | . | Ref. | Ref. | . |
| Currently or in the past[Ŧ] | 1.09 | [0.84,1.43] | 0.519 | 1.11 | [0.84,1.45] | 0.460 | 1.13 | [0.86,1.49] | 0.381 | 1.14 | [0.86,1.50] | 0.355 |

| | OR | (95% CI) | p | OR | (95% CI) | p | OR | (95% CI) | p | OR | (95% CI) | p |
|---|---|---|---|---|---|---|---|---|---|---|---|---|
| Yes, not currently but have happened in the past | 1.04 | [0.75,1.44] | 0.816 | 1.04 | [0.75,1.44] | 0.811 | 1.06 | [0.76,1.46] | 0.742 | 1.06 | [0.76,1.46] | 0.746 |
| Yes, for several years | 0.95 | [0.44,2.08] | 0.902 | 1.01 | [0.46,2.18] | 0.990 | 0.97 | [0.44,2.15] | 0.949 | 1.03 | [0.47,2.25] | 0.948 |
| Yes, for less than a year | 1.73 | [0.89,3.37] | 0.108 | 1.82 | [0.93,3.56] | 0.082 | 1.72 | [0.88,3.38] | 0.113 | 1.81 | [0.92,3.55] | 0.084 |
| Don't want to answer | 0.94 | [0.55,1.61] | 0.818 | 0.96 | [0.61,1.51] | 0.854 | 0.95 | [0.58,1.55] | 0.831 | 0.89 | [0.50,1.61] | 0.712 |
| **Residential area** | | | | | | | | | | | | |
| Urban | Ref. | Ref. | . | Ref. | Ref. | . | Ref. | Ref. | . | Ref. | Ref. | . |
| Suburban | 1.03 | [0.69,1.52] | 0.893 | 0.85 | [0.62,1.17] | 0.324 | 1.04 | [0.70,1.54] | 0.858 | 0.87 | [0.63,1.20] | 0.384 |
| Rural | 0.85 | [0.61,1.18] | 0.328 | 0.99 | [0.67,1.46] | 0.964 | 0.87 | [0.63,1.20] | 0.391 | 1.00 | [0.68,1.49] | 0.991 |
| **Household density** | | | | | | | | | | | | |
| <2 | Ref. | Ref. | . | Ref. | Ref. | . | Ref. | Ref. | . | Ref. | Ref. | . |
| ≥2 | 1.52 | [0.99,2.34] | 0.056 | 1.51 | [0.98,2.32] | 0.060 | 1.57* | [1.02,2.42] | 0.043 | 1.56* | [1.01,2.41] | 0.044 |

| | **(Model 1) Demographic covariates** | | | **(Model 2) Health covariates** | | | **(Model 3) SES covariates** | | | **(Model 4) Fully adjusted model** | | |
|---|---|---|---|---|---|---|---|---|---|---|---|---|
| | OR | (95% CI) | p | OR | (95% CI) | p | OR | (95% CI) | p | OR | (95% CI) | p |
| **SARS-CoV-2 close contact** | | | | | | | | | | | | |
| No | Ref. | Ref. | . | Ref. | Ref. | . | Ref. | Ref. | . | Ref. | Ref. | . |
| Yes | 3.96*** | [2.64,5.94] | 0.000 | 3.94*** | [2.67,5.83] | 0.000 | 3.99*** | [2.65,6.01] | 0.000 | 3.96*** | [2.66,5.88] | 0.000 |
| **Weight status±±** | | | | | | | | | | | | |
| Normal weight | Ref. | Ref. | . | Ref. | Ref. | . | Ref. | Ref. | . | Ref. | Ref. | . |
| Underweight | 1.28 | [0.70,2.35] | 0.423 | 1.32 | [0.72,2.41] | 0.372 | 1.29 | [0.71,2.37] | 0.405 | 1.33 | [0.72,2.43] | 0.362 |
| Overweight-Obese | 1.20 | [0.87,1.65] | 0.261 | 1.17 | [0.85,1.61] | 0.339 | 1.19 | [0.86,1.64] | 0.295 | 1.15 | [0.84,1.59] | 0.388 |
| **Chronic disease±±** | | | | | | | | | | | | |
| No | Ref. | Ref. | . | Ref. | Ref. | . | Ref. | Ref. | . | Ref. | Ref. | . |
| Yes | 1.21 | [0.86,1.71] | 0.281 | 1.19 | [0.85,1.68] | 0.308 | 1.22 | [0.87,1.73] | 0.253 | 1.21 | [0.86,1.71] | 0.279 |
| **Smoking status±±** | | | | | | | | | | | | |
| Never & Former smokers | Ref. | Ref. | . | Ref. | Ref. | . | Ref. | Ref. | . | Ref. | Ref. | . |
| Current smokers | 0.74 | [0.50,1.09] | 0.129 | 0.74 | [0.50,1.09] | 0.123 | 0.75 | [0.51,1.11] | 0.150 | 0.75 | [0.51,1.11] | 0.148 |
| **Blood group±±** | | | | | | | | | | | | |
| O | Ref. | Ref. | . | Ref. | Ref. | . | Ref. | Ref. | . | Ref. | Ref. | . |
| Other | 1.27 | [0.92,1.76] | 0.144 | 1.26 | [0.92,1.74] | 0.151 | 1.24 | [0.90,1.71] | 0.191 | 1.23 | [0.90,1.70] | 0.200 |
| Don't know | 1.07 | [0.72,1.59] | 0.740 | 1.05 | [0.71,1.56] | 0.809 | 1.06 | [0.72,1.58] | 0.761 | 1.05 | [0.70,1.55] | 0.826 |

OR: odds ratio, CI: confidence interval
* $p < 0.05$, ** $p < 0.01$, *** $p < 0.001$
Model 1: adjusted for age only
Model 2: adjusted for age, weight status, chronic disease, smoking status and blood group
Model 3: adjusted for age, education, occupational position and family income
Model 4: adjusted for all the above
± Models 3 and 4 adjusted for the other SES covariates
±± Models 2 and 4 adjusted for the other Health covariates
Ƭ An aggregated analysis with the collapsed categories was conducted first and further disaggregated in the categories below

**Annex IV.** Association between socio-economic and health predictors with SARS-CoV-2 seropositive status, complete models in men

| | (Model 1) Demographic covariates | | | (Model 2) Health covariates | | | (Model 3) SES covariates | | | (Model 4) Fully adjusted model | | |
|---|---|---|---|---|---|---|---|---|---|---|---|---|
| | OR | (95% CI) | p | OR | (95% CI) | p | OR | (95% CI) | p | OR | (95% CI) | p |
| **Education**[±] | | | | | | | | | | | | |
| Tertiary | Ref. | Ref. | . | Ref. | Ref. | . | Ref. | Ref. | . | Ref. | Ref. | . |
| Secondary | 1.15 | [0.64,2.08] | 0.642 | 1.15 | [0.61,2.17] | 0.660 | 1.08 | [0.53,2.19] | 0.831 | 1.07 | [0.51,2.24] | 0.860 |
| Apprenticeship | 0.93 | [0.55,1.58] | 0.795 | 0.84 | [0.47,1.49] | 0.547 | 0.65 | [0.34,1.27] | 0.210 | 0.61 | [0.30,1.23] | 0.169 |
| Compulsory - None | 1.20 | [0.52,2.78] | 0.667 | 1.12 | [0.46,2.72] | 0.802 | 0.97 | [0.35,2.66] | 0.953 | 0.89 | [0.31,2.56] | 0.834 |
| **Occupational position**[±] | | | | | | | | | | | | |
| Higher | Ref. | Ref. | . | Ref. | Ref. | . | Ref. | Ref. | . | Ref. | Ref. | . |
| Lower | 1.70* | [1.02,2.82] | 0.042 | 1.60 | [0.95,2.69] | 0.075 | 1.82 | [1.00,3.32] | 0.052 | 1.79 | [0.97,3.32] | 0.064 |
| Other | 0.65 | [0.26,1.60] | 0.350 | 0.68 | [0.27,1.74] | 0.426 | 0.59 | [0.21,1.67] | 0.325 | 0.65 | [0.22,1.89] | 0.424 |
| **Family income**[±] | | | | | | | | | | | | |
| High | Ref. | Ref. | . | Ref. | Ref. | . | Ref. | Ref. | . | Ref. | Ref. | . |
| Medium | 1.18 | [0.65,2.15] | 0.587 | 1.19 | [0.61,2.24] | 0.593 | 1.14 | [0.60,2.17] | 0.693 | 1.15 | [0.58,2.26] | 0.686 |
| Low | 1.25 | [0.62,2.54] | 0.530 | 1.26 | [0.59,2.77] | 0.547 | 1.14 | [0.52,2.51] | 0.741 | 1.17 | [0.51,2.66] | 0.717 |
| Don't know/No answer/ | 1.25 | [0.61,2.53] | 0.543 | 1.24 | [0.57,2.60] | 0.574 | 1.29 | [0.59,2.85] | 0.525 | 1.29 | [0.57,2.94] | 0.545 |
| **Ethnicity** | | | | | | | | | | | | |
| Caucasian | Ref. | Ref. | . | Ref. | Ref. | . | Ref. | Ref. | . | Ref. | Ref. | . |
| Other | 1.10 | [0.58,2.09] | 0.771 | 1.13 | [0.57,2.23] | 0.734 | 1.05 | [0.52,2.12] | 0.899 | 1.04 | [0.50,2.18] | 0.908 |
| **Country of birth** | | | | | | | | | | | | |
| Switzerland | Ref. | Ref. | . | Ref. | Ref. | . | Ref. | Ref. | . | Ref. | Ref. | . |
| Other HICs | 1.06 | [0.68,1.65] | 0.795 | 1.06 | [0.66,1.69] | 0.805 | 0.97 | [0.59,1.58] | 0.895 | 0.97 | [0.58,1.61] | 0.896 |
| LMICs | 1.03 | [0.52,2.04] | 0.937 | 0.98 | [0.47,2.03] | 0.963 | 0.93 | [0.44,1.96] | 0.844 | 0.86 | [0.39,1.88] | 0.709 |
| **Employment status** | | | | | | | | | | | | |
| Employed | Ref. | Ref. | . | Ref. | Ref. | . | Ref. | Ref. | . | Ref. | Ref. | . |
| Independent | 0.96 | [0.43,2.17] | 0.924 | 1.06 | [0.45,2.48] | 0.902 | 1.02 | [0.43,2.41] | 0.960 | 1.10 | [0.45,2.69] | 0.834 |
| Retired | 0.51 | [0.20,1.32] | 0.167 | 0.52 | [0.20,1.41] | 0.204 | 0.41 | [0.15,1.16] | 0.093 | 0.42 | [0.14,1.21] | 0.107 |
| Student | 0.50 | [0.18,1.35] | 0.170 | 0.56 | [0.20,1.57] | 0.272 | 0.38 | [0.09,1.63] | 0.193 | 0.42 | [0.09,1.91] | 0.263 |
| Unemployed | 0.85 | [0.30,2.42] | 0.759 | 0.84 | [0.28,2.52] | 0.759 | 0.73 | [0.24,2.22] | 0.575 | 0.73 | [0.23,2.33] | 0.595 |
| Other | 0.14 | [0.02,1.14] | 0.066 | 0.12 | [0.01,1.03] | 0.054 | 0.10* | [0.01,0.93] | 0.043 | 0.08* | [0.01,0.85] | 0.036 |
| **Facing financial hardship** | | | | | | | | | | | | |
| Never | Ref. | Ref. | . | Ref. | Ref. | . | Ref. | Ref. | . | Ref. | Ref. | . |
| Currently or in the past[Ͳ] | 1.69* | [1.03,2.78] | 0.037 | 1.62 | [0.97,2.72] | 0.065 | 1.64 | [0.97,2.78] | 0.065 | 1.57 | [0.91,2.70] | 0.103 |

| | | | | | | | | | | | | |
|---|---|---|---|---|---|---|---|---|---|---|---|---|
| Yes, not currently but have happened in the past | 1.66* | [1.00,2.74] | 0.048 | 1.59 | [0.95,2.68] | 0.078 | 1.65 | [0.97,2.79] | 0.065 | 1.58 | [0.92,2.70] | 0.097 |
| Yes, for several years | 1.75 | [0.62,4.93] | 0.288 | 1.75 | [0.58,5.25] | 0.317 | 1.64 | [0.53,5.06] | 0.391 | 1.63 | [0.50,5.29] | 0.418 |
| Yes, for less than a year | 1.54 | [0.53,4.45] | 0.429 | 1.39 | [0.46,4.27] | 0.561 | 1.42 | [0.46,4.41] | 0.546 | 1.25 | [0.38,4.03] | 0.714 |
| Don't want to answer | 0.76 | [0.33,1.73] | 0.509 | 0.69 | [0.29,1.66] | 0.396 | 0.67 | [0.26,1.70] | 0.399 | 0.60 | [0.23,1.60] | 0.308 |
| **Residential area** | | | | | | | | | | | | |
| Urban | Ref. | Ref. | . | Ref. | Ref. | . | Ref. | Ref. | . | Ref. | Ref. | . |
| Suburban | 0.51** | [0.31,0.84] | 0.008 | 0.49* | [0.29,0.84] | 0.009 | 0.51* | [0.30,0.87] | 0.014 | 0.49* | [0.28,0.85] | 0.012 |
| Rural | 0.77 | [0.43,1.38] | 0.379 | 0.75 | [0.40,1.41] | 0.377 | 0.84 | [0.45,1.58] | 0.597 | 0.82 | [0.43,1.59] | 0.558 |
| **Household density** | | | | | | | | | | | | |
| <2 | Ref. | Ref. | . | Ref. | Ref. | . | Ref. | Ref. | . | Ref. | Ref. | . |
| ≥2 | 1.26 | [0.69,2.32] | 0.451 | 1.31 | [0.70,2.46] | 0.404 | 1.18 | [0.62,2.24] | 0.622 | 1.22 | [0.63,2.37] | 0.549 |

| | **(Model 1) Demographic covariates** | | | **(Model 2) Health covariates** | | | **(Model 3) SES covariates** | | | **(Model 4) Fully adjusted model** | | |
|---|---|---|---|---|---|---|---|---|---|---|---|---|
| | OR | (95% CI) | p | OR | (95% CI) | p | OR | (95% CI) | p | OR | (95% CI) | p |
| **SARS-CoV-2 close contact** | | | | | | | | | | | | |
| No | Ref. | Ref. | . | Ref. | Ref. | . | Ref. | Ref. | . | Ref. | Ref. | . |
| Yes | 4.66*** | [2.27,9.60] | 0.000 | 5.33*** | [2.26,12.59] | 0.000 | 5.14*** | [2.12,12.46] | 0.000 | 5.87*** | [2.17,15.92] | 0.001 |
| **Weight status±±** | | | | | | | | | | | | |
| Normal weight | Ref. | Ref. | . | Ref. | Ref. | . | Ref. | Ref. | . | Ref. | Ref. | . |
| Underweight | 0.20 | [0.01,3.43] | 0.266 | 0.20 | [0.01,3.65] | 0.279 | 0.22 | [0.01,4.75] | 0.338 | 0.23 | [0.01,4.97] | 0.347 |
| Overweight-Obese | 1.69* | [1.03,2.76] | 0.036 | 1.72* | [1.04,2.84] | 0.033 | 1.67* | [1.00,2.79] | 0.049 | 1.70* | [1.02,2.86] | 0.044 |
| **Chronic disease±±** | | | | | | | | | | | | |
| No | Ref. | Ref. | . | Ref. | Ref. | . | Ref. | Ref. | . | Ref. | Ref. | . |
| Yes | 0.91 | [0.54,1.52] | 0.711 | 0.87 | [0.50,1.50] | 0.621 | 0.97 | [0.55,1.68] | 0.904 | 0.93 | [0.52,1.66] | 0.798 |
| **Smoking status±±** | | | | | | | | | | | | |
| Never & Former smokers | Ref. | Ref. | . | Ref. | Ref. | . | Ref. | Ref. | . | Ref. | Ref. | . |
| Current smokers | 0.97 | [0.59,1.61] | 0.914 | 0.97 | [0.57,1.72] | 0.899 | 0.96 | [0.55,1.66] | 0.883 | 0.96 | [0.54,1.70] | 0.891 |
| **Blood group±±** | | | | | | | | | | | | |
| O | Ref. | Ref. | . | Ref. | Ref. | . | Ref. | Ref. | . | Ref. | Ref. | . |
| Other | 1.43 | [0.84,2.44] | 0.188 | 1.46 | [0.84,2.55] | 0.183 | 1.43 | [0.81,2.52] | 0.216 | 1.46 | [0.81,2.61] | 0.207 |
| Don’t know | 1.45 | [0.84,2.50] | 0.183 | 1.48 | [0.83,2.61] | 0.180 | 1.40 | [0.79,2.47] | 0.245 | 1.42 | [0.78,2.59] | 0.245 |

OR: odds ratio, CI: confidence interval
* $p < 0.05$, ** $p < 0.01$, *** $p < 0.001$
Model 1: adjusted for age only
Model 2: adjusted for age, weight status, chronic disease, smoking status and blood group
Model 3: adjusted for age, education, occupational position and family income
Model 4: adjusted for all the above
± Models 3 and 4 adjusted for the other SES covariates
±± Models 2 and 4 adjusted for the other Health covariates
Ƭ An aggregated analysis with the collapsed categories was conducted first and further disaggregated in the categories below

**This supplementary material belongs to the main paper:** Socio-economic determinants of SARS-CoV-2 infection: results from a population-based serosurvey in Geneva, Switzerland

[ii] Office cantonal de la statistique (OCSTAT) Genève. Revenus et fortune des ménages. Available at : https://www.ge.ch/statistique/domaines/20/20_02/tableaux.asp

[iii] World Bank Country and Lending Groups – World Bank Data Help Desk [Internet]. [cited 10 nov 2021]. Available at: https://datahelpdesk.worldbank.org/knowledgebase/articles/906519-world-bank-country-and-lending-groups

[iv] Richard A, Wisniak A, Perez-Saez J, Garrison-Desany H, Petrovic D, Piumatti G, et al. Seroprevalence of anti-SARS-CoV-2 IgG antibodies, risk factors for infection and associated symptoms in Geneva, Switzerland: a population-based study: Scand J Public Health [Internet]. 19 oct 2021 [cited 10 nov 2021]; Available at: https://journals.sagepub.com/doi/full/10.1177/14034948211048050

[v] Office cantonal de la statistique (OCSTAT) Genève. Population du Canton de Genève selon l’origine et le statut migratoire. Communications statistiques No 55 – Avril 2017. Available at : https://www.ge.ch/statistique/tel/publications/2017/analyses/communications/an-cs-2017-55.pdf

[vi] Office cantonal de la statistique (OCSTAT) Genève. Population résidante selon l'origine, le sexe et le plus haut niveau de formation achevée. Available at : https://www.ge.ch/statistique/domaines/15/15_03/tableaux.asp

[vii] Office cantonal de la statistique (OCSTAT) Genève. Répartition de la population résidante active occupée âgée de 15 ans ou plus selon diverses caractéristiques. Available at : https://www.ge.ch/statistique/domaines/03/03_02/tableaux.asp

[viii] Zufferey J. La santé dans le canton de Genève: résultats de l'Enquête suisse sur la santé 2017. Observatoire Suisse de la Santé; 2020

[ix] Perelli I. Adéquation des groupes sanguins entre donneurs et patients genevois (Doctoral dissertation, University of Geneva). 2020
